# The source of individual heterogeneity shapes infectious disease outbreaks

**DOI:** 10.1101/2021.02.18.21251983

**Authors:** Baptiste Elie, Christian Selinger, Samuel Alizon

**Affiliations:** MIVEGEC, Univ Montpellier, CNRS, IRD, Montpellier, France; Swiss Tropical and Public Health Institute, Basel, Kreuzstrasse 2, 4123 Allschwil, Switzerland; Center for Interdisciplinary Research in Biology (CIRB), Collège de France, CNRS, INSERM, Université PSL, Paris, France

**Keywords:** epidemiology, modelling, infection duration, superspreading, evolutionary rescue, emerging infectious diseases

## Abstract

There is known heterogeneity between individuals in infectious disease transmission patterns. The source of this heterogeneity is thought to affect epidemiological dynamics but studies tend not to control for the overall heterogeneity in the number of secondary cases caused by an infection. To explore the role of individual variation in infection duration and transmission rate on parasite emergence and spread, while controlling for this potential bias, we simulate stochastic outbreaks with and without parasite evolution. As expected, heterogeneity in the number of secondary cases decreases the probability of outbreak emergence. Furthermore, for epidemics that do emerge, assuming more realistic infection duration distributions leads to faster outbreaks and higher epidemic peaks. When parasites require adaptive mutations to cause large epidemics, the impact of heterogeneity depends on the underlying evolutionary model. If emergence relies on within-host evolution, decreasing the infection duration variance decreases the probability of emergence. These results underline the importance of accounting for realistic distributions of infection rates to anticipate the effect of individual heterogeneity on epidemiological dynamics.

---

The expected number of secondary cases produced by an infected individual in a naive population is a key concept in epidemiology [6, 32]. It is classically referred to as the basic reproduction number and denoted *R*_0_. Only infections with *R*_0_ > 1 can cause major outbreaks. However, this mean value does not reflect the impact of super-spreading events, where an individual causes an unusually large number of secondary cases [19, 27, 36, 40, 44, 58]. The more frequent these events are, the higher the variance in the number of secondary cases, and, therefore, the lower the probability of outbreak emergence and the faster the epidemic growth for outbreaks that do emerge [40].

Several biological processes can explain the heterogeneity in the number of secondary cases [53]. However, models investigating these processes tend only to vary one source of heterogeneity at a time. By doing so, they do not control for the (overall) heterogeneity in the number of secondary cases, which is known to have strong effects, independently of its source [40]. One of the few exceptions suggests that the biology matters since it finds, for instance, that heterogeneity in host susceptibility has a lesser impact on the probability of emergence than heterogeneity in transmission rate, which can be defined as the product between a contact rate and the probability of transmission given that there is a contact between two individuals [59].

We use a stochastic mechanistic model to explore whether heterogeneity in transmission rates or recovery rates have different effects on an epidemic spread. Based on earlier models, we hypothesise that a more homogeneous distribution of infectious period duration decreases the variability of population dynamics in the early outbreak, therefore increasing the probability of outbreak extinction [5], but also increasing epidemic growth as well as epidemic peak size [5, 43]. However, we stress that these hypotheses are based on studies that, contrarily to us, do not control for variations in the distribution of the number of secondary cases.

Even if initially maladapted (*i*.*e. R*_0_ < 1), a parasite can evolve into a well-adapted strain before fading out and then cause a major outbreak, a phenomenon called ‘evolutionary emergence’ or ‘evolutionary rescue’ [8, 23]. Since higher epidemic sizes can be reached more frequently with increasing heterogeneity secondary cases when *R*_0_ < 1 [25], we hypothesise that the source of heterogeneity could affect evolutionary emergence. Since we do not explicitly model the within-host evolution process, we consider two extreme evolutionary processes for a mutant strain with *R*_0_ > 1 to appear [2, 23]: either by taking over a host infected by the resident strain or during a transmission event.

Following earlier studies [25, 29, 40], we assume that the number of secondary infections caused by each individual follows a Negative-Binomial distribution 𝒵 with mean *R*_0_ and dispersion parameter *k*. The smaller *k*, the more dispersed 𝒵. For example, the 2003 SARS outbreak in Singapore led to many superspreading events and transmission chain analyses estimated that *k* = 0.16 [40] and recent data from COVID-19 epidemics yielded values of *k* in the order of 0.3 [51].

We model individual transmission rates and infection duration values using lognormal distributions, denoted respectively ℬ and Γ. Most models involving ordinary differential equations are ‘memoryless’ -that is the duration of the infections is assumed to be exponentially distributed (*CV*_Γ_ = 1) (but see [5, 39, 43]). This is biologically unrealistic for recovery events since they often depend on the number of days since infection [15, 37] and tends to overestimate the heterogeneity due to infection duration. We disentangle the specific role of infection duration heterogeneity from that of the secondary cases by varying *k*, and the coefficient of variation (CV) of the infection duration (*CV*_Γ_). Those two parameters combined govern the distribution of transmission rate.

We simulate outbreaks, without and with evolution, and measure key summary statistics to analyze the impact of different sources of heterogeneity on emerging outbreaks properties. We confirm that the dispersion of the distribution of the number of secondary infections (𝒵) is the main driver of the frequency of emergence, but we also find that the source of this heterogeneity has a strong impact on the properties of emerging epidemics, and more interestingly that it can affect the risk of evolutionary emergence.

As an illustration, we compare dynamics that could be obtained with parameters estimated from two outbreaks: SARS in Singapore in 2003 and Ebola in West Africa in 2014, which have similar values of *R*_0_ and *k* [4, 40] and different infection duration heterogeneity. We estimate *CV*_Γ_ = 1.04 (95 % credible interval (CI): 0.44-1.9) for Ebola and 0.27 (95 % CI: 0.01 - 0.80) for SARS. An explanation for that difference is that the Ebola virus is known to sometimes persist in some body fluids after clearance from the blood [16]. Animal studies also show variability in the host immune response against Ebolavirus infection, which might allow persistence for some individuals [42]. Regarding SARS outbreaks, the reason why some infected individuals spread more than others the virus is thought to be a combination of host and environmental properties. On the biological side, individuals causing superspreading events were older [50], and coinfections have been hypothetised to increase the infectivity of SARS-CoV [11]. On the environmental side, superspreaders had a higher number of close contacts, and the diagnosis of the infection was often delayed [50].

## Material and Methods

### Model without evolution

We implement a non-Markovian version of the Susceptible-Infected-Recovered (SIR) epidemiological model [33], which means that not all rates are held constant throughout an infection [26]. We assume that the host population is of fixed size *N* and that epidemics are initiated by a single infectious individual. At time *t*, each individual is characterized by its current state (susceptible, infectious, or removed), and, if infected, the time at which it will recover.

The first source of heterogeneity in the model comes from the transmission rates and has a behavioural (i.e. contact rates) or a biological (i.e. infectiousness) origin. We model it by drawing the *per capita* transmission rate *β*_*i*_ for each individual *i* from a lognormal distribution, denoted ℬ, with parameters *μ*_*B*_ and *σ*_*B*_. For mathematical convenience, and without further qualitative impact, we set the mean of ℬ such that 𝔼 [*B*] *N* = 1. The standard deviation of ℬ is imposed by the choice of the coefficient of variation (*CV*_*B*_) which is equal 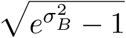.

The second source of heterogeneity comes from the infection duration and has a biological origin. We assume that individuals remain in the *I* compartment for a time drawn randomly from a lognormal distribution, noted Γ, with parameters *μ*_Γ_ and *σ*_Γ_. By construction, the expectation of Γ is *R*_0_ in our model and we vary its coefficient of variation, which is equal to 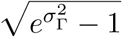, between 0.05 and 2.

### Coefficients of variation and 𝒵 dispersion

Given the construction of our model, the distribution od the number of secondary infections (𝒵) is determined by heterogeneities in transmission rate and infection duration. Since the force of infection over the course of an individual’s infection is the product of two lognormal distributions (ℬ and Γ), it is itself log-normally distributed, with parameters *μ*_*Z*_ = *μ*_*B*_ + *μ*_Γ_ and 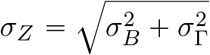. 𝒵 is therefore a lognormal-Poisson compound distribution.

### Evolutionary emergence model

We introduce an additional class of individuals by distinguishing between *I*_*r*_ and *I*_*m*_, which refer to individuals infected by the resident (resp. mutant) parasite strain, with reproduction number 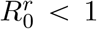 (resp. 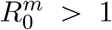). Initially, we assume that *I*_*r*_ = 1 and *I*_*m*_ = 0. Mutant infections can emerge from a transmission event or from taking over an infected host. In the case of within-host mutation, the mutation rate represents the instantaneous probability that a mutant appears and takes over the host. In the case of mutation during transmission, it represents the probability that a mutant is transmitted instead of a resident strain. We assume that the mutation increases the mean transmission rate without altering *CV*_*B*_ (*i*.*e*. by setting 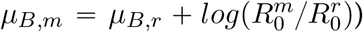. We further assume that the infectious period duration is not impacted by the mutation. For simplicity, we neglect coinfections and therefore assume that, in the case of within-host mutations, the mutant instantaneously takes over the host.

### Frequency of emergence

We use the total epidemic size to determine if an outbreak has emerged or not. Emergence is assumed to occur when the total epidemic size is greater than the herd immunity threshold, *i*.*e*. 1 −1/*R*_0_ [6].

### Numerical simulations

We simulate epidemics, i.e. the succession of infection and recovery events, using Gillespie’s next reaction method [26] to generate non-Markovian distributions. The algorithm runs as follows:

1. *Initialize (i*.*e. set S, I* = 1, *t* = 0)
2. *In case of new infected individual i, draw β*_*i*_ *and the recovery time of this individual from the distribution* Γ.
3. *Update the new force of infection* 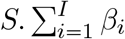 *and draw the time to the next infection assuming an exponential distribution*.
4. *Look for the event with the closest time of occurrence (i*.*e. either recovery or new infection), and update the compartments (S, I)*.
5. *Update the time t to the time of the new event*.
6. *Go back to step 2*.

In case of evolutionary emergence, we adapt the model in function of the way the mutant appears. i) If the mutant appears during transmission, the model includes one force of infection for each class of infected host (*I*_*r*_ and *I*_*m*_), and two additional events: infection by the mutant strain (assuming an exponential distribution with a rate 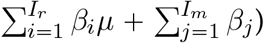, and recovery of an *I*_*m*_ individual. ii) In the scenario where the mutant first takes over the host, we distinguish the event of infection by the mutant strain (assuming an exponential distribution with a rate 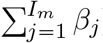) from the within-host mutation of a resident strain into a mutant strain (assuming an exponential distribution with a rate *I*_*r*_ × *μ*).

The model was implemented in Java 11.0.7 using parallel computation to decrease computing time. Simulation outputs were analyzed with R v.4.1.2. The scripts used are available at https://gitlab.in2p3.fr/ete/heterogeneity-outbreak.

### Parameters estimation for known outbreaks

To estimate *CV*_*B*_ and *CV*_Γ_ from observed outbreaks, we analyzed serial interval and secondary cases distributions from Measles [31], Ebola [20, 57], pneumonic plague [24], Smallpox [21, 46], Monkeypox [30] and SARS outbreaks [38, 40]. For the Measles outbreak, patient line data were available, therefore allowing joint distribution estimations, and for the others, we had to assume that the two distributions were independent (see the Table S1 for further details about the data and parameters sources).

To obtain biologically relevant parameters from these empirical data, we infer parameters assuming a model with a latent period, the distribution of which we set using independent sources in the literature [9, 35, 45, 47, 57]. For simplicity, we assume that for a given parasite the distribution of the latent period does not vary between outbreaks. We also use independent estimates of *R*_0_ [31, 38, 57]. We also assume a constant transmission rate during infectious period. We use a Bayesian approach, with the following priors: *CV*_*B*_ ∼ 𝒩 (2, 10) and *CV*_Γ_ ∼ 𝒩 (0.5, 1). We use jags v. 4.3.0 to estimate parameters.

## Results

### Epidemics emergence without evolution

For a given secondary cases heterogeneity *k*, the coefficients of variation in infection duration (*CV*_Γ_) and transmission rate (*CV*_*B*_) are negatively correlated. This is shown in Figure 1 and further explained in the Methods. The former being easier to measure, we focus on the role of infection duration heterogeneity, but the results can also be interpreted in terms of infectivity heterogeneity.

**Figure 1:**
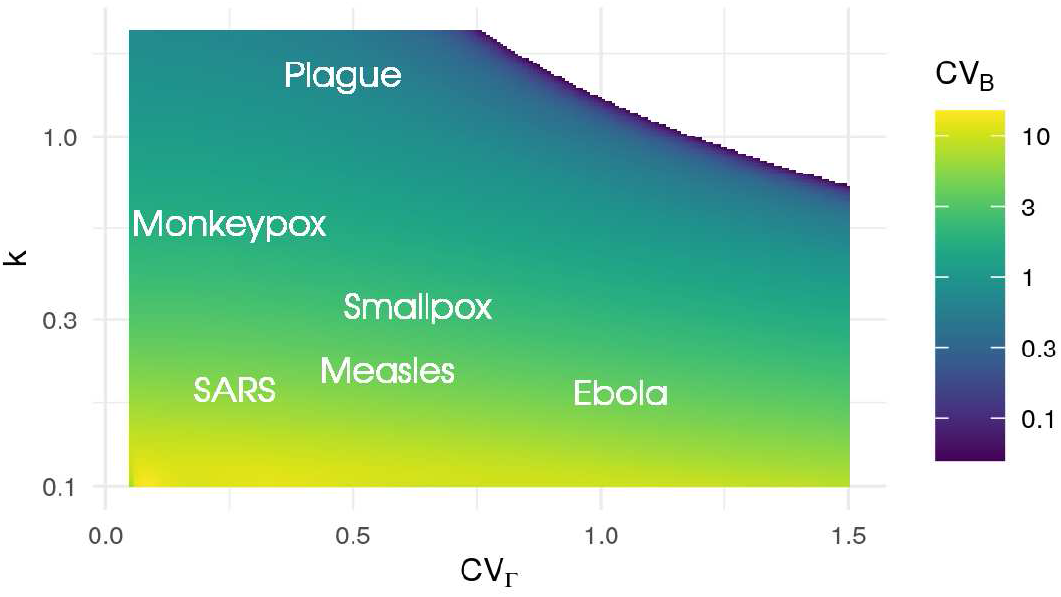
Numerical estimation of the infection rate coefficient of variation (*CV*_*B*_), as a function of *k* and *CV*_Γ_. Names in white show the range of values estimated using maximum likelihood methods from outbreak data. If *k* remains constant, increasing *CV*_Γ_ always decreases *CV*_*B*_. Note that when secondary cases heterogeneity is low (*i*.*e. k* is high) it is impossible to have a high *CV*_Γ_.

To illustrate the feasibility to infer these infection properties, we highlight the parameter value for several well-studied outbreaks in Figure 1. This also shows that our parameters ranges are biologically realistic.

### Probability of emergence

Figure 2A shows that the probability of an outbreak emergence only depends on the overall 𝒵 heterogeneity, here measured by *k*. The source of heterogeneity *(i*.*e*. infection duration or infectiousness) does not seem to play any role. Results are shown with *R*_0_ = 1.5, but a similar pattern is observed for any *R*_0_ < 1.

**Figure 2:**
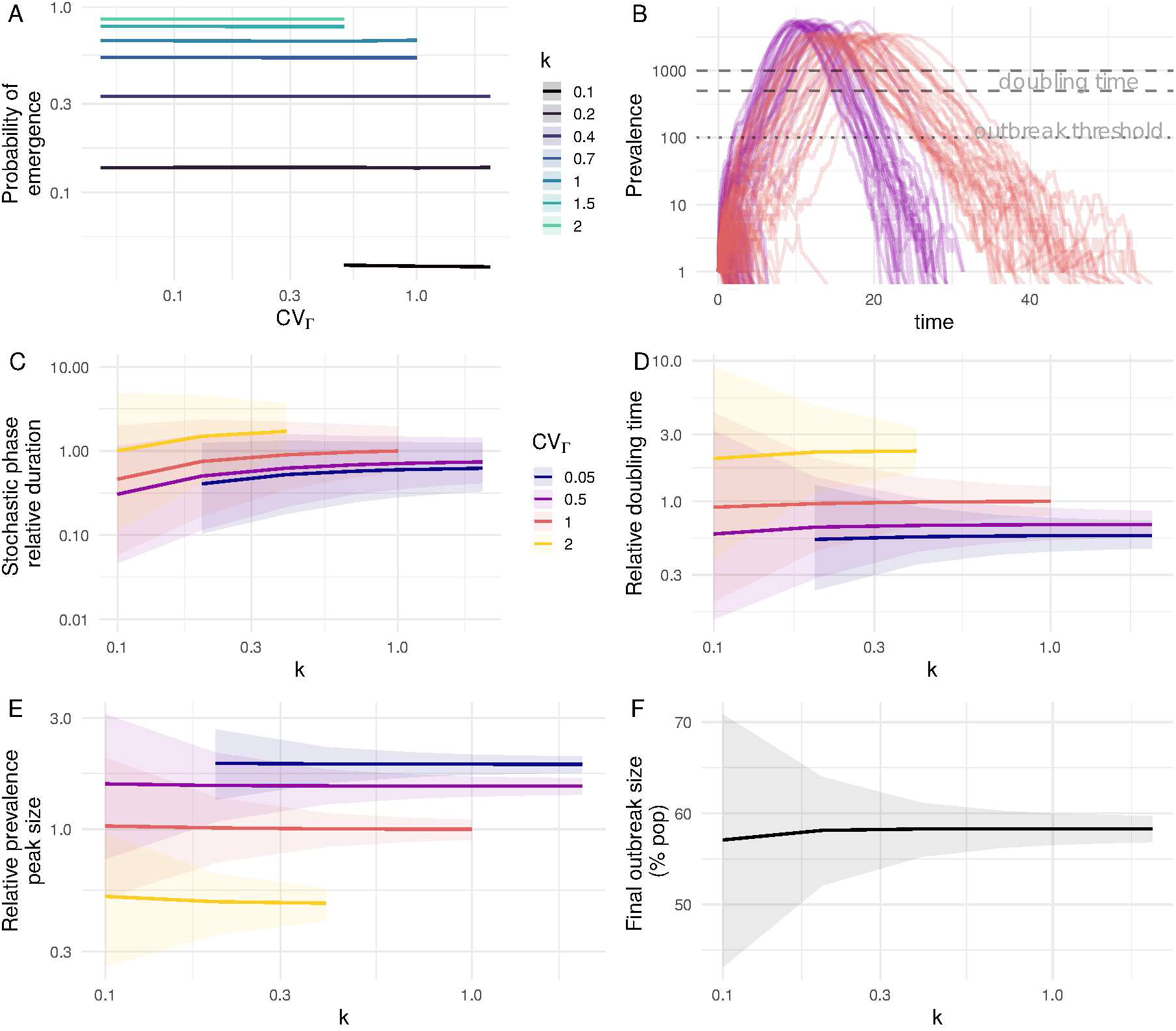
Summary statistics of epidemics emergence without evolution. A) Frequency of emergence of an outbreak starting from one infection as a function of model heterogeneity, with *R*_0_ = 1.5. B) Epidemic trajectories with the same secondary cases distribution *k* = 0.4, but different infection duration heterogeneity (*CV*_Γ_ = 1 in brown and *CV*_Γ_ = 0.5 in red). The total population size is 50,000. C) Relative time until the epidemic reaches the emergence threshold (*i*.*e*. here a prevalence of 100 infected individuals). D) Relative doubling time during the exponential phase (*i*.*e*. going from a prevalence of 500 to 1000 infections). E) Relative prevalence peak size. F) Final outbreak size, as a percentage of the total population. Panels C, D, and E show metrics relative to the case where *k* = 1 and *CV*_Γ_ = 1, and colors indicate the value of *CV*_Γ_. Lines represent mean values computed from simulated outbreaks that emerged and shaded areas the 95% confidence interval.

In the following, we analyze the properties of simulated outbreaks without evolution with *R*_0_ = 1.5 and compare key metrics to a reference value close to the markovian case, *i*.*e. k* = 1 and *CV*_Γ_ = 1.

### Growth rate

In the initial phases of an outbreak, the law of large numbers does not apply and prevalence time series shown are strongly affected by stochasticity (Fig. 2B). We quantify the early growth during this stochastic phase by measuring the time until the prevalence reaches the outbreak threshold of 100 infected individuals [28]. As expected [40], decreasing *k* leads to faster epidemic growth. Furthermore, for a given *k*, increasing the heterogeneity in infection duration also increases the early epidemic growth (Fig. 2C). On average, this would make a SARS outbreak reach the outbreak threshold 50% faster than an Ebola outbreak.

We then study the deterministic exponential growth phase, which starts when the number of infected is high enough to reach the law of large numbers, and ends when the depletion of susceptible host population cannot be neglected anymore [28] (Fig. 2B). Figure 2D shows that the growth rate during this phase is mostly impacted by *CV*_Γ_. For instance, even with similar *R*_0_, Ebola outbreaks would have a doubling time of 1.4 times the mean infection duration, while SARS outbreaks would have a doubling time of 0.9 time the mean infection duration. Not taking into account the difference in infectious period distribution between the two epidemics and considering a memoryless model with *CV*_Γ_ =)1 would lead to an overestimation of the SARS *R*_0_ [55].

### Epidemic peak size and final size

The prevalence peak value is highly affected by the heterogeneity in infection duration: its median increases by more than 50% when *CV*_Γ_ decreases from 1 to 0.5 (Fig. 2 E). *k* has little effect on the mean epidemic peak size, but there is a correlation between the variance in peak size and that of 𝒵.

Finally, none of our heterogeneity metric seems to affect the median final epidemic size, which is always close to 58% of the population (Fig. 2 F), corresponding to the expected value for *R*_0_ = 1.5 according to classical theory [33]. As for the other metrics, the variance in the total epidemic size decreases with *k*.

### Evolutionary emergence

We now assume that the introduced ‘resident’ strain has a 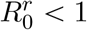 and, therefore, will go extinct unless it evolves into a phenotypically different ‘mutant’ strain with 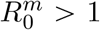. The mutant strain can arise either by taking over a host infected by the resident strain or during a transmission event.

### Mutation probability

To disentangle the evolutionary process from the epidemiological process, following Yates *et al*.[59], we first assume that a mutant instantaneously takes over the population 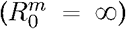. The probability of mutant emergence does not depend on the origin of heterogeneity. Moreover, figures 3A and B show that the way the mutant strain appears does not seem to affect the relationship between the frequency of emergence and the secondary cases heterogeneity *k*: when 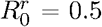, there is little impact of *k* on the frequency of emergence, whereas when 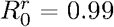, increasing *k* increases the frequency of emergence.

**Figure 3:**
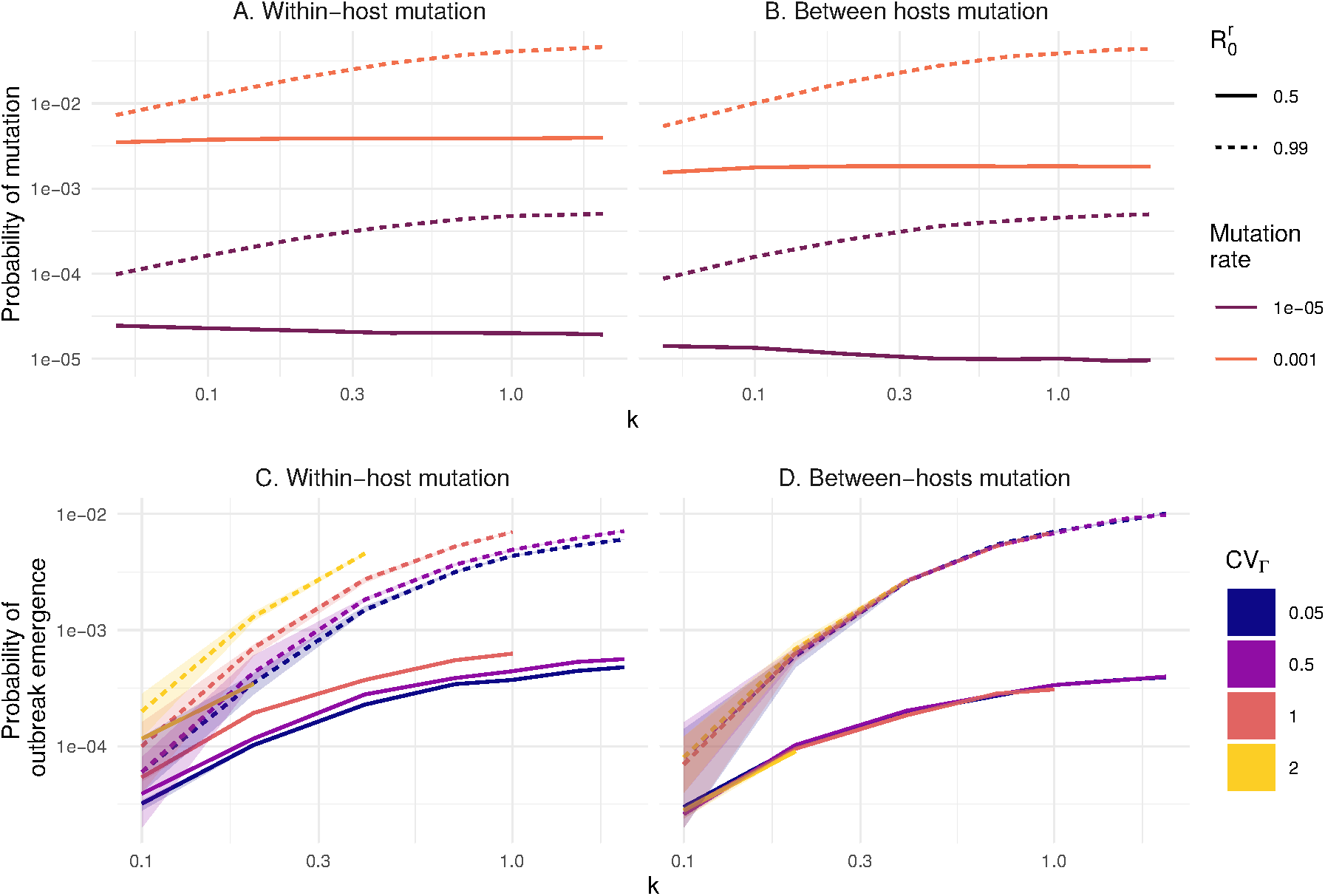
Individual heterogeneity and evolutionary emergence. Probability of mutation, as a function of the mutation origin, mutation rate and 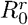. A) the mutant appears during the infection within a host and replaces the resident strain, or B) the mutant appears during transmission. Since there is no difference in function of *CV*_Γ_, the results shown combine the whole range of *CV*_Γ_. Probability of outbreak emergence of a mutant C) taking over a host and D) appearing during transmission, in the case 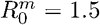 and a mutation rate of 10^−3^.

### Mutant outbreak

We then consider the more realistic case where the mutant has a *R*_0_ = 1.5 (Fig. 3C, D). The general trend is qualitatively consistent with the case without evolution: decreasing the secondary cases heterogeneity increases the frequency of emergence.

When the mutant appears during transmission (Fig. 3D), the source of heterogeneity does not play any role. However, when the mutant appears by taking over an infection (Fig. 3C), decreasing the infection duration heterogeneity increases the probability of emergence. The difference between these two scenarios is that when the mutant arises within the host, the infection is ongoing, and the host recovery time is kept constant since we assume no difference in immune response between resident and variant strains. Therefore, with a more heterogeneous infection duration, individuals with longer infections will increase the probability that a mutant arises within the host and can transmit before the host recovery.

## Discussion

When modeling epidemics, the variation between individuals can be aggregated into a single metric, the dispersion of the secondary infections caused by each individual, which shapes infectious diseases outbreaks [40]. Several studies investigate how variations in a specific trait can have an impact on epidemiological dynamics but the majority overlook that variations in one trait (e.g. the distribution of the duration of infectious periods) may also affect the distribution of individual secondary cases. In this study, we investigate the relative effects of variation in the infection duration and transmission rate while keeping the distribution of the secondary cases constant.

Increasing the heterogeneity in transmission rates is known to lead to a faster increase in cases per generation among the outbreaks that do emerge in branching process models [40]. By simulating the whole course of the epidemic, we show that this effect does not translate into an increased growth rate after the epidemic evades the stochastic phase. Methodologically, this could also be studied using recent developments of branching process theory in epidemiology to incorporate the depletion of susceptible hosts [10].

We show that the heterogeneity in infectious period duration plays an important role in the deterministic phase of the epidemic, by increasing the growth rate and, more strikingly, the prevalence peak size. While previous studies reported a similar effect on both secondary cases heterogeneity and infection duration heterogeneity [5, 13, 55, 56], we further show that this phenomenon is intrinsically related to the latter. Indeed, more heterogeneous infectious periods are known to lead to longer generation times because transmission relies on long infections, therefore increasing the doubling time and flattening the epidemic curve [55].

When considering a simple evolutionary rescue scenario, we show that the probability of mutation does not depend on the infection duration heterogeneity. This is consistent with the observation that the final epidemic size is not affected by the source of heterogeneity. Furthermore, we show that with very low 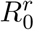, secondary cases distribution does not have any impact either. This can be explained by the fact that for *R*_0_ < 1, the decrease in frequency of emergence associated with heterogeneity is compensated by a higher probability in reaching larger outbreak sizes [25] (Fig. S1), therefore maintaining the mean outbreak size (Fig. S2). This effect diminishes as *R*_0_ gets higher and disappears when *R*_0_ > 1.

Finally, we show that infectious period duration heterogeneity can affect evolutionary emergence depending on the process that generates the mutant infection [23]. The impact of the mutational pathway and evolutionary scenario has already been pointed out by several studies [1, 59]. As expected, we find no difference between the two mutation scenarios if the process is memoryless. This further underlines the importance of questioning this biologically-unrealistic assumption [5, 13, 39, 43]. When assuming more realistic infection duration distributions, we find that if mutations appear upon transmission events, the probability of evolutionary emergence only depends on the distribution of the secondary cases. However, when the mutation appears after a host takeover, infection duration heterogeneity increases the frequency of emergence. This is illustrated by figure S4: although in either scenario the probability that a mutant appears remains constant and equal to the mutation rate, when the mutation occurs within the host, the probability that it gets transmitted is higher in the case of rare long infections, as already pointed out [7].

Our effort to maintain a simple and tractable model of outbreak emergence naturally leads to several limitations. In particular, there is an identifiability issue regarding the biological bases of the transmission rate heterogeneity, which could originate from variations in transmission rate or in host susceptibility. However, Yates *et al*. [59] find that the heterogeneity in infectivity plays a larger role in the frequency of emergence than the heterogeneity in susceptibility. It could also be interesting to enrich the model by considering a latent period during which exposed hosts are not yet infectious. This has been shown to affect *R*_0_ estimates but in a deterministic model that did not take into account superspreading events [56]. More generally, investigating other sources of heterogeneity of the number of secondary infections may help uncover potential biases. Another simplification made here is the assumption that infectiousness is constant over the infectious period of and individual. This is biologically not true, and therefore the infectious period defined here is probably shorter than the real infectious period, since infectiousness is usually higher at the beginning of the epidemic.

Since we ignore within-host dynamics, we chose two extreme scenarios regarding the way a mutant appears: either during transmission or within the host. Biological reality is likely in-between: mutants will gradually take over a host, which means an increasing proportion of the transmission events will be caused by the mutant [2]. At least for rapidly evolving viruses such as HIV-1 and HCV, within-host genetic variation is higher than what is expected given the strong host immune response selection [49]. This shows that within-host selection of novel mutations and transmission occur at the same rate.

Nested models, which explicitly include both within and between-host dynamics, can take into account this gradual replacement. Coombs *et alii* [17] showed in a simple nested model with chronic infections that the best between-hosts competitors can be competitively excluded if they are outcompeted within the host in the short term during an infection. Moreover, when allowing mutation, coexistence of both strains could be possible under certain scenarios, which was not possible with our simplified model. When taking explicitly into account the interaction between the parasite and the host’s immune system and the possibility of multiple infections, models suggest that the outcome of the competition can lead to the coexistence of two strains with different within-host growth rates, as soon as there is a possibility that multiple infections can occur [3]. Including the possibility that more than two strains can coexist during the infection, it was shown that the level of selection that matters depends on the extent of phenotypic variation: with a higher between-host than within-host phenotypic variation is observed, it is expected that strains maximizing the between-host transmission are selected, and vice-versa [41]. Finally, Park *et alii* [48] combined a nested model with the question of the probability of emergence of an outbreak, with a stochastic epidemiological model. They showed that conflicting fitness effects of a mutation at the within-host level and at the between-host levels can strongly decrease the probability of emergence of a mutant.

We assumed that the population has no spatial structure, which is more realistic for directly transmitted diseases, such as SARS or measles, than for sexually transmitted infections for which contact networks impose strong constraints [25]. Furthermore, at the beginning of an epidemic, the spatial structure appears to have little effect on outbreak metrics, especially *R*_0_ [52]. However, it is known that heterogeneity in host susceptibility and spatial structure decrease the final epidemic size, *i*.*e*. the total proportion of the population infected throughout the epidemic [12, 54]. We also do not include host demography and limit our analysis to a single epidemic wave.

We also assumed no correlation between infectiousness and infectious period duration. While this seems biologically realistic, little is known about the nature of the relationship between those parameters. Indeed, one could expect that higher infectiousness is associated with a higher pathogen load, leading to a shorter asymptomatic period where transmission can occur, as it has been observed for HIV infections [22]. However, when analyzing the Measles outbreak from Hagelloch, where a joint estimation of both parameters is possible, we found no significant correlation between the estimated infectious rate and the infectious period duration (Fig. S3), although our sample is limited to the 32 individuals who did transmit early in the outbreak.

Finally, this analysis relies on numerical results. This enables us to explore the role of stochasticity, which is particularly important to consider in the context of outbreak emergence from a mathematical modelling [14] and a statistical inference [34] point of view. However, it limits our analysis to the area of punctual parameters that we selected as being biologically relevant.

These theoretical results have implications for outbreak monitoring. In particular, we show that making simplifying but biologically unrealistic assumptions about the distributions of infection duration can lead to underestimating the risk of emergence, the epidemic doubling time, and the prevalence peak size. Given the risk of saturation of healthcare systems, accurately anticipating these values is a major issue. This stresses the importance of collecting detailed biological data to better inform epidemiological models.

## Data Availability

code is available on gitlab: https://gitlab.in2p3.fr/ete/heterogeneity-outbreak/

## Acknowledgements

The authors thank the CNRS, the IRD, and acknowledge the itrop HPC (South Green Platform) at IRD Montpellier for providing HPC resources that have contributed to the research results reported within this study (https://bioinfo.ird.fr/).

This paper has been submitted on a preprint server [18].

