## Supplementary material for "The source of individual heterogeneity shapes infectious disease outbreaks": Supplemantary figures

February 4, 2022

### Parameters estimation for known outbreaks

A limitation of the estimations of $CV_{\Gamma}$ and $CV_B$ from real epidemics is that we could only find joint distributions of secondary cases and serial interval in a measles outbreak. For the others, we used independent distributions for the other outbreaks. However, although this assumption does increase the confidence interval, we do not expect it to bias our results because any potential correlation is expected to be minimal. For instance, in the measles outbreak we analysed, the Spearman correlation coefficient between serial interval and secondary cases generated by the infector was 0.20.

Moreover, assuming constant infectiousness during the infectious period and absence of infection during the latent period, we infer the mean infection duration as $\bar{\Gamma} = 2(\bar{s} - \bar{e})$, where $\bar{s}$ is the mean serial interval and $\bar{e}$ is the mean latent period.

| Outbreak | Secondary cases | Serial interval | $R_0$ | Latent period | $CV_B$ (95% CI) | $CV_\Gamma$ (95% CI) | mean $\Gamma$ (days) | Note |
|---|---|---|---|---|---|---|---|---|
| SARS Singapore | data from 57 cases [9] | Data from 180 cases [8] | 1.63 [9] | Gamma (mean 5.2d, sd 2.5d) [7] | 5.93 (3.08 - 9.82) | 0.36 (0.03 - 0.89) | | Serial interval and incubation period distribution cover the whole outbreak / secondary cases represent only the early phase before public health interventions. |
| Measles Hagelloch | Data from 66 cases | [6] | 2.2 (from secondary cases data) | Gamma (mean 8.58d, sd 1.33 d) [1] | 4.4 (1.90 - 8.3) | 0.69 (0.12 - 1.71) | 3.64 | Outbreak in a German village in 1865. Data were restricted to the first third of the transmission events. Incubation period estimated from an independent outbreak (264 families from Providence, Rhode Island). |
| Ebola Guinea | Data from 152 individuals [2] | Data from 192 individuals [13] | 1.71 [13] | Data from 155 individuals [13] | 4.10 (1.83-7.26) | 1.04 (0.45 - 2.00) | 10.1 | Transmission chains come from Guinea, whereas the other data come from the same outbreak but in the whole region of West Africa. The whole outbreak is considered: heterogeneity in infection duration might be affected by public health measures. |
| Smallpox in Europe | 32 independent importations - only the first indigenous generation [3] | 5 outbreaks in England and India [11] (n=223) | 3.19 (from secondary cases data) | 131 cases [10] | 3.3 (1.45 - 6.62) | 0.71 (0.52-0.96) | 6.93 | The dataset for latent period, secondary cases, and serial intervals come from different outbreaks. |
| Pneumonic Plague | Data from 74 cases (6 outbreaks) | [4] | Inferred: 1.32 [4] | | 0.67 | 0.48 | 2.5 | In this case, we directly used the infection period duration coefficient of variation estimated in the study and retrieved $CV_B$ from the secondary cases heterogeneity. No confidence interval is available with this computation. |
| Monkeypox | 147 cases in Zaïre 1980-1984 [5] | 62 cases in Zaïre 1980-1984 [5] | 0.32 (from secondary cases data) | 28 cases, 2013 outbreak in the Dem. Rep. of Congo [12] | 1.83 (0.72-3.80) | 12.7 | 0.12 (0.02-0.32) | The data for incubation period and secondary cases come from the same geographic region, but 30 year apart. |

Table S1: Parameters, data sources for each studied outbreak.

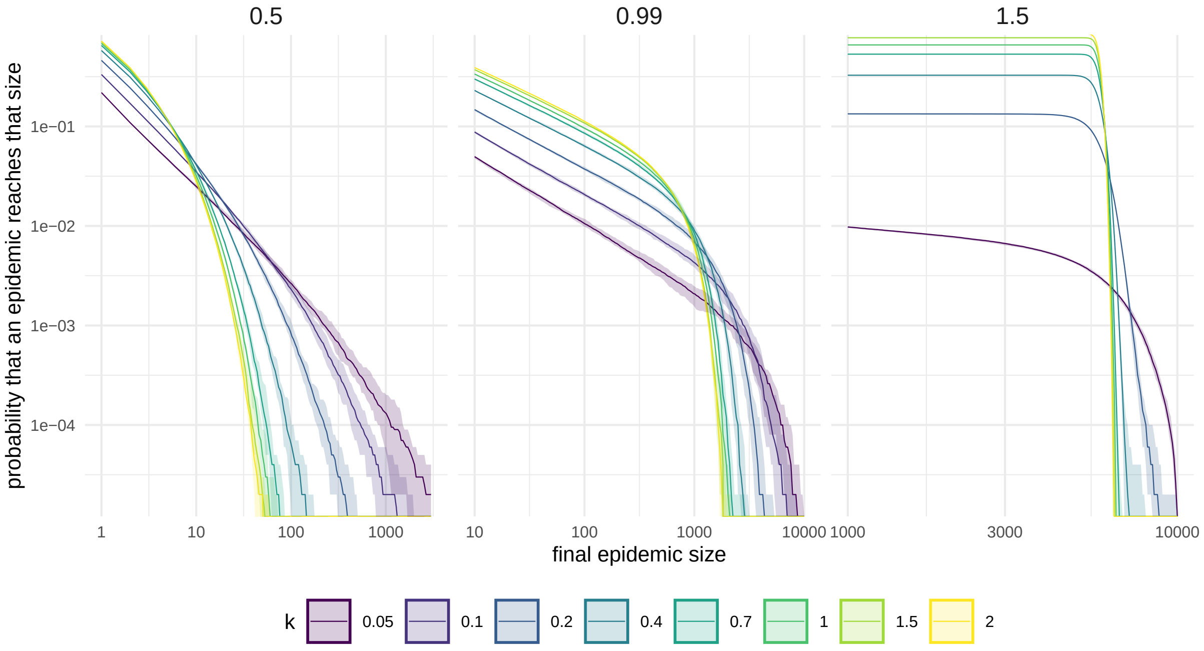


Figure S1: **Final epidemic size cumulative distribution, as a function of secondary cases heterogeneity.** Results are presented in the case without evolution, and with three different $R_0$: 0.5, 0.99, 1.5. No difference was observed between the sources of heterogeneity, and the results are presented combining all ranges of simulated $CV_\Gamma$.

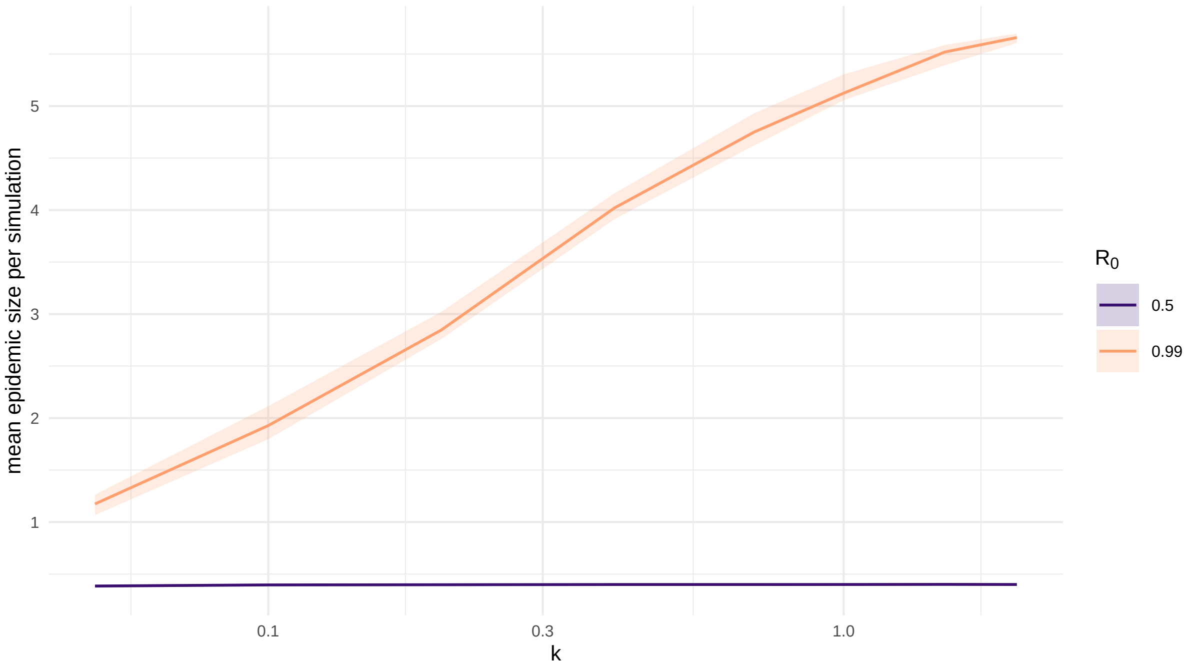


Figure S2: **Mean number of infected individuals during an epidemic, as a function of the secondary cases heterogeneity and $R_0$.**

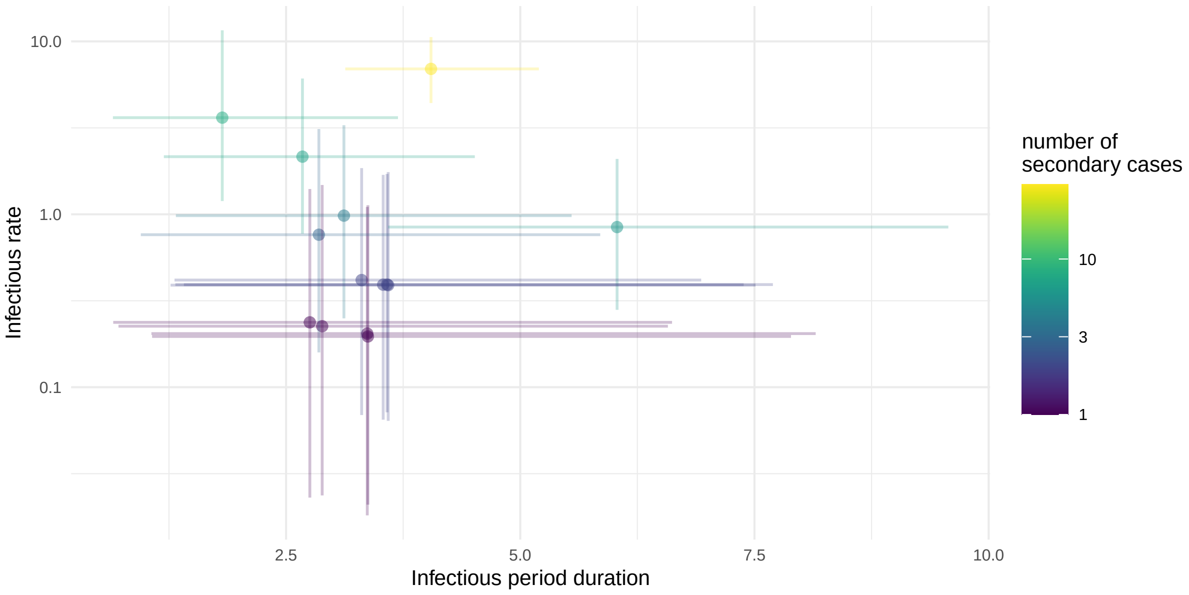


Figure S3: **Relationship between the estimated transmission rate and infection duration for Measles outbreak in Hagelloch.** The parameters could be jointly estimated thanks to patient line data [6] in our Bayesian model. No significant correlation was found between the two metrics, even when removing the superspreading event (Spearman's rank correlation $\rho = -0.34$, $p = 0.25$). Line ranges represent the 95% credible interval.

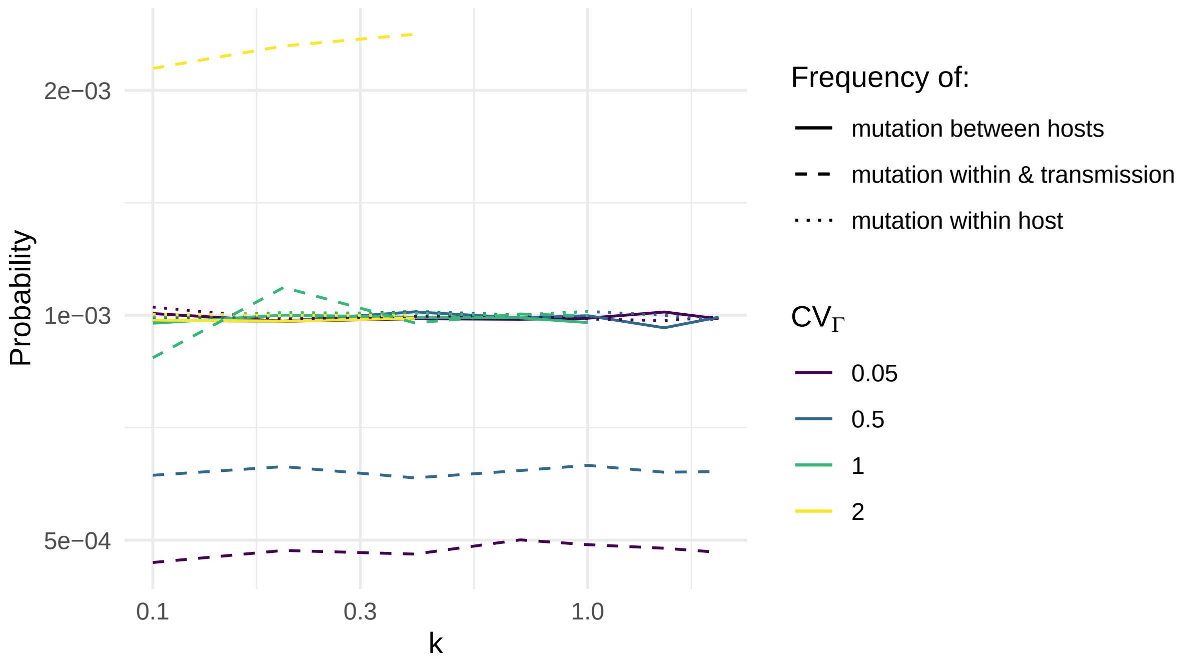


Figure S4: **Probability of mutation as a function of the mutation scenario, $k$ and $CV_\Gamma$.**

r0_range.pdf

Figure S5: **Sensitivity analysis: the impact of $R_0$ on the metrics without evolution.** In order to simplify the reading, results are shown for only two values of $CV_\Gamma$. Results should be read similarly to figure **??**. For each $R_0$ value, relative results are shown with $k = 1$, $CV_\Gamma = 1$ and the corresponding $R_0$ as a reference. A. Frequency of emergence as a function of k and $R_0$. B. Stochastic phase relative duration. C. Relative doubling time. D. Prevalence peak size. E. Final outbreak size.
